# Type 1 Diabetes Risk Phenotypes Using Cluster Analysis

**DOI:** 10.1101/2023.10.10.23296375

**Authors:** Lu You, Lauric A. Ferrat, Richard A. Oram, Hemang M. Parikh, Andrea K. Steck, Jeffrey Krischer, Maria J. Redondo, Type 1 Diabetes TrialNet Study Group

## Abstract

**Background:** Although statistical models for predicting type 1 diabetes risk have been developed, approaches that reveal clinically meaningful clusters in the at-risk population and allow for non-linear relationships between predictors are lacking. We aimed to identify and characterize clusters of islet autoantibody-positive individuals that share similar characteristics and type 1 diabetes risk.

**Methods:** We tested a novel outcome-guided clustering method in initially non-diabetic autoantibody-positive relatives of individuals with type 1 diabetes, using the TrialNet Pathway to Prevention (PTP) study data (n=1127). The outcome of the analysis was time to type 1 diabetes and variables in the model included demographics, genetics, metabolic factors and islet autoantibodies. An independent dataset (Diabetes Prevention Trial of Type 1 Diabetes, DPT-1 study) (n=704) was used for validation.

**Findings:** The analysis revealed 8 clusters with varying type 1 diabetes risks, categorized into three groups. Group A had three clusters with high glucose levels and high risk. Group B included four clusters with elevated autoantibody titers. Group C had three lower-risk clusters with lower autoantibody titers and glucose levels. Within the groups, the clusters exhibit variations in characteristics such as glucose levels, C-peptide levels, age, and genetic risk. A decision rule for assigning individuals to clusters was developed. The validation dataset confirms that the clusters can identify individuals with similar characteristics.

**Interpretation:** Demographic, metabolic, immunological, and genetic markers can be used to identify clusters of distinctive characteristics and different risks of progression to type 1 diabetes among autoantibody-positive individuals with a family history of type 1 diabetes. The results also revealed the heterogeneity in the population and complex interactions between variables.

**Funding:** National Institute of Diabetes and Digestive and Kidney Diseases (R01DK121843), the National Institute of Allergy and Infectious Diseases, the Eunice Kennedy Shriver National Institute of Child Health and Human Development (cooperative agreements U01 DK061010, U01 DK061034, U01 DK061042, U01 DK061058, U01 DK085461, U01 DK085465, U01 DK085466, U01 DK085476, U01 DK085499, U01 DK085509, U01 DK103180, U01 DK103153, U01 DK103266, U01 DK103282, U01 DK106984, U01 DK106994, U01 DK107013, U01 DK107014, UC4 DK106993, UC4DK117009), and the Juvenile Diabetes Research Foundation.

**Research in Context:** *Evidence Before This Study:* We searched PubMed on June 12, 2023, using the keywords “cluster type 1 diabetes”, “heterogeneity type 1 diabetes”, and “endotypes type 1 diabetes” with no restrictions on publication date. Only articles published in English were considered. Existing literature suggests that individuals at risk of type 1 diabetes form a diverse population influenced by a combination of genetic and environmental factors. However, there is a scarcity of research focusing on defining clusters within this at-risk population; previous studies using clustering analysis to define diabetes subtypes have primarily utilized unsupervised clustering methods, which may not be as effective in capturing the critical variables that inform disease risks.

*Added Value of This Study:* We applied an outcome-guided clustering analysis to unravel the complexity and heterogeneity of type 1 diabetes. This study introduces a new method for identifying clusters of individuals based on key risk factors and the observed disease outcomes. Within each cluster, individuals will exhibit similar characteristics and diabetes risk, while the clusters themselves represent distinct levels of risk. Unlike previous approaches that employ regression models relying on linearity and additivity assumptions, this method provides advantages by uncovering underlying interactions and correlations among risk factors. Our results were validated in the DPT-1 study cohort.

*Implications of All the Available Evidence:* Our findings suggest that demographic, metabolic, immunological, and genetic markers can be used to identify distinct clusters with varying risks of progression to type 1 diabetes within autoantibody-positive individuals with a family history of the disease. The results demonstrate how different combinations of these risk factors contribute to the risk and how clusters of similar risks can exhibit unique characteristics with regard to the risk factors, which highlights the heterogeneity in this population.

## Introduction

Type 1 diabetes is a chronic disease characterized by immune-mediated destruction of insulin-producing beta cells in the pancreas^1^. Progression to type 1 diabetes is a complicated process that involves interactions between multiple genetic and environmental factors, leading to immunological dysfunction and metabolic abnormalities^2^. Heterogeneity in the causes and processes contributes to explaining the high degree of variation in the risk and rate of progression to clinical disease within individuals at risk for type 1 diabetes^3^. Although previous studies have identified major risk factors^4,5^ and developed models for predicting type 1 diabetes risk^6–10^, there is a need to further understand the heterogeneity of the disease by exploring various levels and patterns of the risk factors in relation to one another and how they contribute to the risks associated with the disease. A majority of the prediction models in the literature used regression models to quantify risk for type 1 diabetes by a linear combination of several selected risk factors.^6–10^ These models mainly focus on the prediction of outcomes and are usually constrained by model assumptions such as linearity and proportionality of hazard rates, that is, the supposition that predictors proportionally affect the risk along the entire spectrum of variation. Therefore, such models are usually not flexible enough to capture the complex interactions between risk factors and thus, may overlook the heterogeneity of the population.

The objective of this study is to use an outcome-guided clustering method to identify groups of individuals with comparable characteristics informed by their type 1 diabetes risks. The study will focus on autoantibody-positive relatives of people with type 1 diabetes. There is a lack of literature on defining clusters among individuals at risk of type 1 diabetes, and previous studies on clustering analysis of diabetes subtypes are limited to unsupervised clustering methods which may be less efficient in capturing the key variables that inform disease risks^11^. The proposed method is a nonparametric approach (meaning that it does rely on assumptions about the data distribution or linear relationships) for identifying clusters of individuals informed by several key risk factors ascertained from the data. Individuals within the same cluster will share similar characteristics and diabetes risk while the clusters correspond to different risks. This method differs from previous attempts to measure risk using regression models^6,7^ in that it does not rely on stringent distributional assumptions to describe the relationship between predictors and outcomes. Instead, the proposed method is more advantageous to allow for relationships that change along the spectrum of the characteristics under study and reveal underlying interactions and correlations between risk factors to further define the etiopathogenesis of type 1 diabetes.

## Materials and Methods

### Population

The TrialNet Pathway to Prevention (PTP) study is a prospective cohort study that follows participants who have a family member with type 1 diabetes and at least one positive islet autoantibody. Enrolled participants are routinely monitored for islet autoantibodies and metabolic status, including hemoglobin A1c (HbA1c) and oral glucose tolerance tests (OGTT). This study included 1127 individuals from the TrialNet PTP study without diabetes at baseline and who were genotyped on the ImmunoChip array, which is an Illumina Infinium genotyping chip designed to study variants in genes known to be associated with autoimmune diseases^12^ (Supplementary Figure S1). The individuals were enrolled between 2004 to 2012 and followed for a median of 2·7 years. To validate the results from the analysis, we used a dataset of 704 initially non-diabetic autoantibody positive participants from the Diabetes Prevention Trial of Type 1 Diabetes (DPT-1) study.

### Data Collection

The primary endpoint in this analysis is the time from baseline OGTT visits to the diagnosis of clinical (stage 3) type 1 diabetes. The last follow-up date is defined as the date of type 1 diabetes diagnosis or, for those who did not progress to diabetes, the last OGTT test. Demographic variables in the TrialNet PTP data included gender, ethnicity, relationship to proband with type 1 diabetes, age and body mass index (BMI) z-scores at baseline visit. BMI z-scores for participants under 20 were calculated using the Centers for Disease Control and Prevention (CDC) growth charts while for participants over 20 years of age the formula for 20-year-olds was used. Islet autoantibodies include glutamic acid decarboxylase autoantibodies (GADA), islet antigen-2 autoantibodies (IA2A), autoantibodies to insulin (mIAA), and islet cell antibodies (ICA).^13^

GADA and IA2A titers from non-harmonized assays^14^ were transformed to the scale of the harmonized ones by the constrained nonparametric B-spline regression method in the R package “cobs”^15^. Metabolic functions were assessed by OGTT and HbA1c tests. The metabolic variables considered are fasting glucose levels, fasting C-peptide levels, the area under the 2-hour glucose response curve (glucose AUC), the area under the 2-hour C-peptide response curve (C-peptide AUC), and HbA1c. Genotype information was collected based on ImmunoChip data^12^. Genetic risk factors in the analysis include the type 1 diabetes genetic risk score-2 (GRS2)^16^, 8 SNPs that were shown to be associated with islet autoantibody positivity by Törn et al.^17^, and the 5 most susceptible human leukocyte antigen (HLA) alleles discovered in the Type 1 Diabetes Genetics Consortium^18^ (labeled S1 to S5) (see Supplementary Tables 1 and 2 for details). The validation dataset from the DPT-1 study contains demographic variables (gender, ethnicity, relationship to proband with type 1 diabetes, age, and BMI z-scores), islet autoantibody titers data (GADA, IA2A, mIAA, and ICA), and metabolic variables (fasting glucose, fasting C-peptide, glucose AUC, and C-peptide AUC) listed above, but no ImmunoChip genotype data is available in the DPT-1 population.

### Statistical Analysis

The descriptive analysis summarizes the distribution of continuous variables by median and interquartile range (IQR), and categorical variables by counts and proportions.

A statistical machine learning method developed by the authors is applied to the clustering analysis of the data. It is an outcome-guided clustering method that can identify clusters of individuals with varying levels of type 1 diabetes risk while maintaining similarity in terms of their characteristics within each cluster. Details about the clustering method is provided in the Appendix with a graphic illustration in Supplementary Figure S2. This approach ensures that the identified clusters are differentiated by their disease risk, while maintaining internal homogeneity within each cluster. As a result, the method can fulfill the goal of identifying clusters of individuals with similar risks but disparate disease risks across clusters. After the clusters are identified, data and variables were visualized in a heatmap organized by their cluster membership, and disease risk in each cluster is assessed by Kaplan-Meier curves. To examine the relationship and closeness of the clusters, we used the multi-dimensional scaling (MDS) method^19^ to map the clusters to a two-dimensional space. The distributions of variables are summarized by medians, IQR and boxplots. To examine how the risks of clusters align with the established risk scores, we summarized the distributions of risk scores (e.g., Diabetes Prevention Trial Risk Score^6^ [DPTRS], and Index60^7^) by clusters. Pseudo R-squared statistics are used to examine the variation in the survival outcomes explained by the clusters as well as the risk scores. We applied the classification tree algorithm in the R package “rpart” to find a simple tree-structured decision rule for assigning new individuals to specific clusters. Finally, we used the DPT-1 cohort to validate the decision rule. Boxplots and Kaplan-Meier curves are used to compare the distribution of variables and type 1 diabetes risk of the clusters in the training and testing datasets.

## Results

We conducted clustering analysis of 1127 initially non-diabetic TrialNet participants with available genetic data. A total of 173 individuals were excluded as their cluster membership cannot be determined due to missing data (Supplementary Figure S1). The baseline characteristics of the 954 individuals with cluster assignment are presented in Table 1 (second column). The results from hierarchical clustering can be visualized in a tree-structured dendrogram (Column 1 of Figure 1). The average Silhouette width^20^ that measures the similarity of individuals within a cluster is maximized at three clusters, with another peak at eight clusters. Other measures to determine the number of clusters indicate that the optimal number of clusters is either three or eight (Supplementary Figure S4), suggesting the possibility of a higher-order structure to the clustering. We analyzed the results at two levels, with 8 clusters categorized into 3 groups. We labeled the three groups by A (with cluster A1), B (with clusters B1, B2, B3 and B4), and C (with clusters C1, C2, and C3) (Figure 1, Column 6). Heatmaps were used to visualize the values of variables and their cluster membership (Figure 1, Columns 2-6). Risk of progression to type 1 diabetes is represented by the Kaplan-Meier curves in Column 7 of Figure 1 and Figure 2 (outer). The estimated diabetes-free survival probability in 1-5 years for each cluster is given in Table 2. The spatial distribution of the 8 clusters on the MDS map is shown in Column 8 of Figure 1 and Figure 2 (center). In Figure 2, we used radar plots to display the distribution of the variables in each cluster. The medians and IQR of variables by cluster are summarized in Supplementary Table S4.

**Figure 1.**
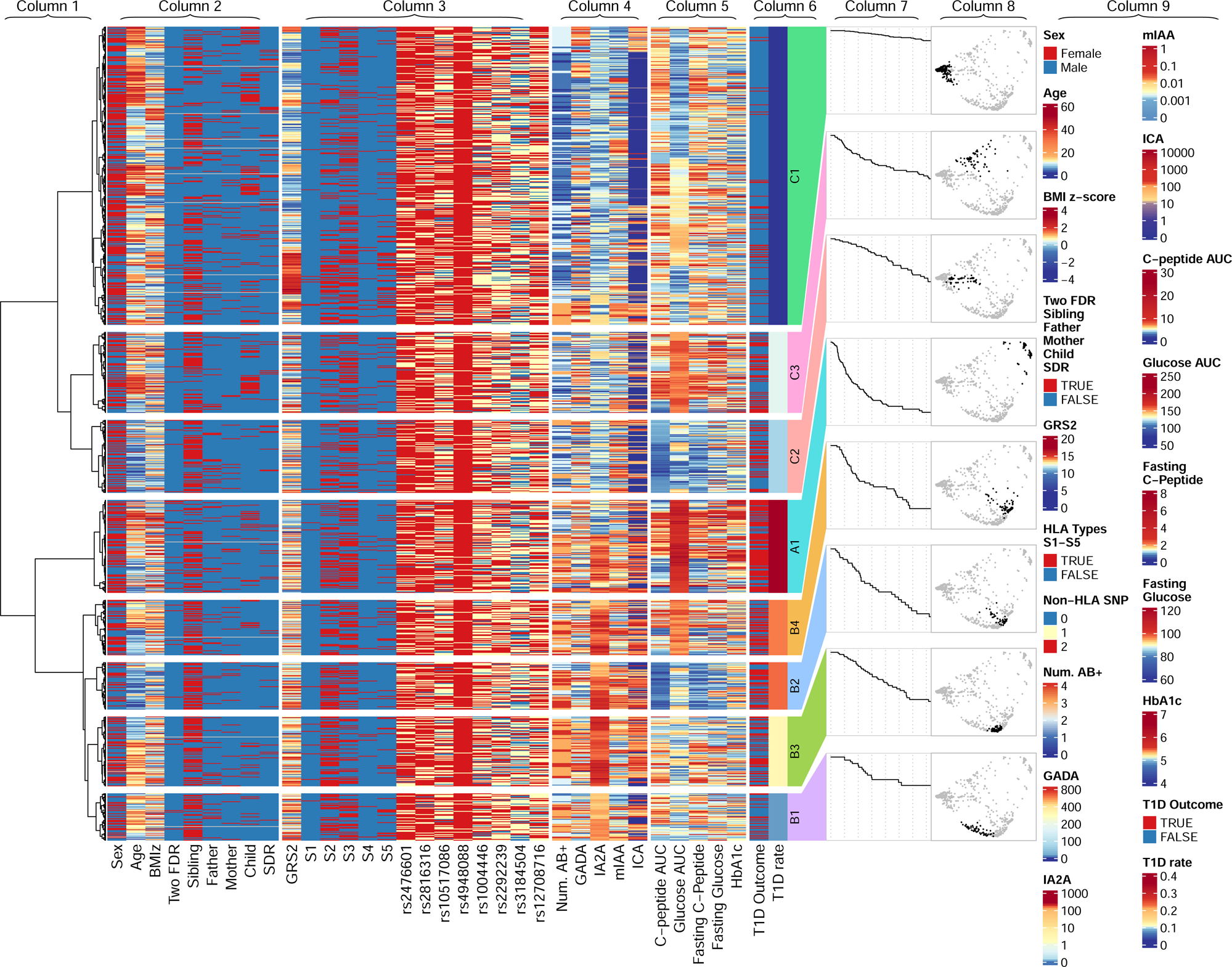
Dendrogram and heatmap resulted from the clustering analysis. From left to right, the displayed information is the complete-linkage dendrogram (Column 1), the heatmap of demographic information (Column 2), the heatmap of genetic information (Column 3), the heatmap of immunological markers (Column 4), the heatmap of metabolic markers (Column 5), the heatmap of T1D outcomes and rates (Column 6), diabetes-free Kaplan-Meier survival curves of clusters (Column 7), distribution of clusters on the multidimensional scaling map (Column 8), and, in the foremost right side, the legends denoting the color scheme for the variables (Column 9).

**Figure 2.**
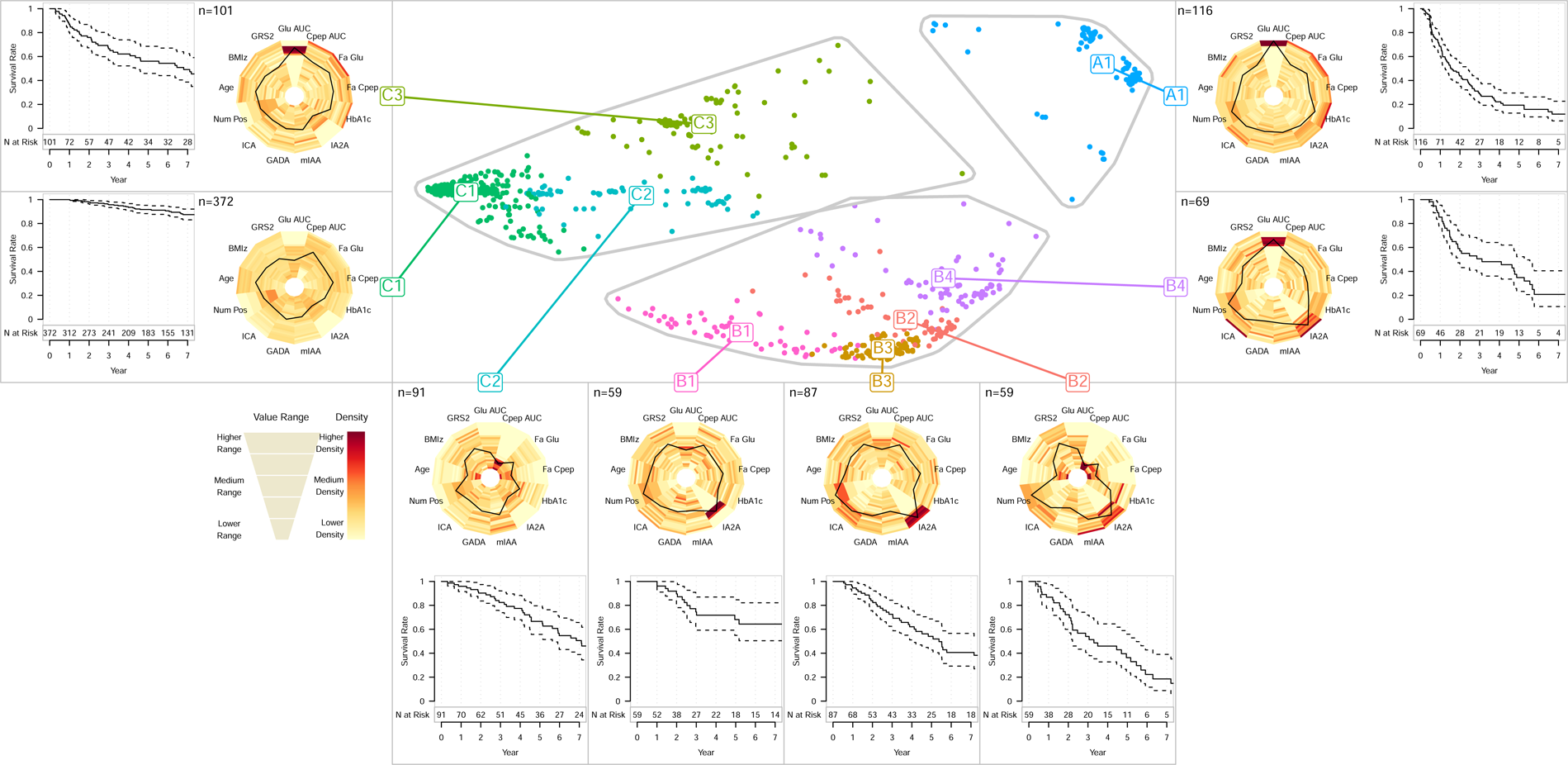
MDS map of clusters (center), flanked by the radar charts of each cluster (surrounding the MDS maps), and the Kaplan-Meier survival curves by cluster with their corresponding 95% confidence intervals (outer).

**Table 1.** Baseline characteristics of TrialNet PTP participants and DPT-1 participants included in the analysis. Numbers in the table are count (frequency) for categorical variables and median (IQR) for continuous variables.

| Variable | TrialNet PTP | DPT-1 |
| --- | --- | --- |
| N | 954 (100.0%) | 706 (100.0%) |
| Sex | .. | .. |
| Female | 503 (52.9%) | 311 (44.1%) |
| Male | 448 (47.1%) | 395 (55.9%) |
| Ethnicity | .. | .. |
| Hispanic or Latino | 77 (8.3%) | 30 (4.2%) |
| Not Hispanic or Latino | 846 (91.7%) | 676 (95.8%) |
| Relatives with Type 1 Diabetes | .. | .. |
| Child | 151 (16.0%) | 184 (26.1%) |
| Parent | 183 (19.4%) | 31 (4.4%) |
| Sibling | 488 (51.8%) | 385 (54.5%) |
| Two or More FDR† | 48 (5.1%) | 47 (6.7%) |
| SDR* | 72 (7.6%) | 59 (8.4%) |
| BMI-z Score | 0.54 (-0.22,1.21) | 0.89 (0.24,1.54) |
| Age (Years) | 11 (7,21) | 11 (8,16) |
| Number of Positive Autoantibodies | 2 (1,3) | 3 (2,3) |
| GADA Titer (DK units/ml) | 282 (30,575) | 475 (39,695) |
| IA2A Titer (DK units/ml) | 1.6 (0.1,198.0) | 9 (0,262) |
| mIAA Titer (index) | 0.005 (0.001,0.022) | 0.001 (0.000,0.013) |
| ICA Titer (JDF units) | 0 (0,80) | 80 (40,320) |
| GADA Positivity | 716 (75.1%) | 518 (73.4%) |
| IA2A Positivity | 418 (43.8%) | 372 (52.7%) |
| mIAA Positivity | 360 (37.7%) | 174 (24.6%) |
| ICA Positivity | 444 (46.5%) | 703 (99.6%) |
| Glucose AUC (mg/dl) | 129 (115,148) | 128 (113,147) |
| Fasting Glucose (mg/dl) | 89 (84,95) | 86 (80,92) |
| C-peptide AUC (pmol/ml) | 4.9 (3.7,6.6) | 3.7 (2.7,4.7) |
| Fasting C-peptide (pmol/ml) | 1.4 (1.0,1.9) | 0.9 (0.6,1.3) |
| HbA1c (%) | 5.1 (4.9,5.3) | 5.3 (5.1,5.6) |
| Type 1 Diabetes GRS2 | 12.5 (11.0,13.8) | Not Available |

\*SDR: second-degree relative(s)
†FDR: first-degree relative(s)

**Table 2.** The diabetes-free survival probability of the 8 identified clusters. Values in the parentheses are the corresponding 95% CI.

| Cluster | N | 1-Year Survival | 2-Year Survival | 3-Year Survival | 4-Year Survival | 5-Year Survival |
| --- | --- | --- | --- | --- | --- | --- |
| A1 | 116 | 0.69 (0.60,0.78) | 0.44 (0.35,0.55) | 0.29 (0.21,0.40) | 0.22 (0.15,0.32) | 0.19 (0.13,0.30) |
| B1 | 59 | 1.00 (1.00,1.00) | 0.90 (0.81,0.99) | 0.74 (0.62,0.89) | 0.72 (0.59,0.87) | 0.68 (0.55,0.85) |
| B2 | 59 | 0.87 (0.78,0.97) | 0.72 (0.60,0.87) | 0.54 (0.41,0.72) | 0.46 (0.33,0.64) | 0.36 (0.24,0.56) |
| B3 | 87 | 0.94 (0.89,1.00) | 0.84 (0.76,0.93) | 0.73 (0.62,0.84) | 0.62 (0.51,0.76) | 0.54 (0.43,0.69) |
| B4 | 69 | 0.85 (0.76,0.95) | 0.57 (0.45,0.72) | 0.51 (0.39,0.66) | 0.48 (0.36,0.64) | 0.35 (0.23,0.52) |
| C1 | 372 | 1.00 (1.00,1.00) | 0.98 (0.96,1.00) | 0.96 (0.93,0.98) | 0.94 (0.91,0.97) | 0.92 (0.88,0.95) |
| C2 | 91 | 0.96 (0.92,1.00) | 0.90 (0.84,0.97) | 0.82 (0.74,0.92) | 0.77 (0.68,0.88) | 0.67 (0.56,0.80) |
| C3 | 101 | 0.87 (0.80,0.95) | 0.76 (0.67,0.86) | 0.66 (0.57,0.78) | 0.62 (0.52,0.74) | 0.56 (0.46,0.69) |

Group A consists of one cluster A1 with the highest risk (2-year diabetes-free survival rate 0·44 [95% CI: 0·35,0·55]). From Figure 1 and Supplementary Table S4, we can observe that this cluster has the highest glucose AUC, fasting glucose, and HbA1c.

Group B consists of four clusters B1, B2, B3, and B4 (2-year diabetes-free survival rate, B1: 0.90 [95% CI: 0·81,0·99], B2: 0·72 [95% CI: 0·60,0·87], B3: 0·84 [95% CI: 0·76,0·93], B4: 0·57 [95% CI: 0·45,0·72]). These clusters are characterized by increased autoantibody titers, especially in IA2A titer and ICA titer, and numbers of positive autoantibodies, with additional distinctive characteristics that are unique to each of them. Cluster B4 represents individuals with high glucose AUC. Cluster B2 represents individuals with younger ages, lower glucose AUC, lower C-peptide AUC, lower fasting C-peptide. The remaining two clusters B1 and B3 have similar metabolic characteristics in terms of glucose AUC, fasting glucose, C-peptide AUC, and fasting C-peptide, but their immunological characteristics are quite different. Cluster B3 represents individuals with increased IA2A titer, ICA titer, and number of autoantibodies.

Cluster B1 is a lower risk cluster with older age and lower IA2A titer. (See Figure 1 and Supplementary Table S4.)

Group C consists of three clusters C1, C2, and C3 with lower glucose and autoantibody titers (2-year diabetes-free survival rate, C1: 0·98 [95% CI: 0·96-1·00], C2: 0·90 [95% CI: 0·84-0·97], C3: 0·76 [95% CI: 0·67-0·86]). Cluster C1 is the cluster with the lowest risk, characterized by older age, lower glucose AUC, higher C-peptide AUC, lower GRS2, and lower IA2A titer. Compared to Cluster C1, Cluster C2 had an increased risk characterized by younger age, lower C-peptide AUC, lower fasting C-peptide, slightly higher GRS2, and higher number of positive autoantibodies. Cluster C3 had the highest risk among the three clusters with higher glucose AUC and higher fasting glucose. (See Figure 1 and Supplementary Table S4.)

To compare our results with established risk scores in the literature, in Supplementary Figure S3, we present the distributions of DPTRS^6^ and Index60^7^ by cluster. DPTRS is a risk score derived from OGTT measures, BMI, and age. Index60 is another risk score that combines OGTT-derived glucose and C-peptide values. Both DPTRS and Index60 have been found to correlate well with disease risks^7,21–23^. Supplementary Figure S3 shows that there is a reasonable alignment between DPTRS, Index60, and disease risks, where higher risk clusters generally have higher DPTRS and Index60. We also fitted three separate proportional hazards models with DPTRS, Index60, and the cluster membership being the covariates respectively, and compared the pseudo R-squared values^24^ of the three model fits. The pseudo R-squared values are 0·30 (95% CI: 0·25-0·35), 0·22 (95% CI: 0·17-0·27), and 0·28 (95% CI: 0·23-0·32) respectively, which suggests that the variability in the disease outcome explained by the cluster information is comparable to the DPTRS and Index60. We note that while DPTRS and Index60 are continuous risk scores used to predict disease outcomes, the cluster information offers a unique perspective on the heterogeneity of the disease by identifying different combinations and levels of risk factors that contribute to similar risk of developing T1D, resulting in distinct groups of individuals.

Last, we applied the classification tree algorithm to find a decision rule to assign individuals to one of the 8 identified clusters. As shown in Figure 3, glucose AUC, IA2A titers and C-peptide AUC are the primary variables for assigning individuals to the clusters. Individuals with glucose AUC higher than 162 mg/dl are assigned to Clusters A1. Individuals with glucose AUC between 144 and 162 mg/dl are assigned to clusters B4 or C3 based on IA2A titers. Individuals with glucose AUC under 144 are classified based on IA2 titers and C-peptide AUC. The cutoff values were determined based on the points that most effectively distinguish the clusters.

**Figure 3.**
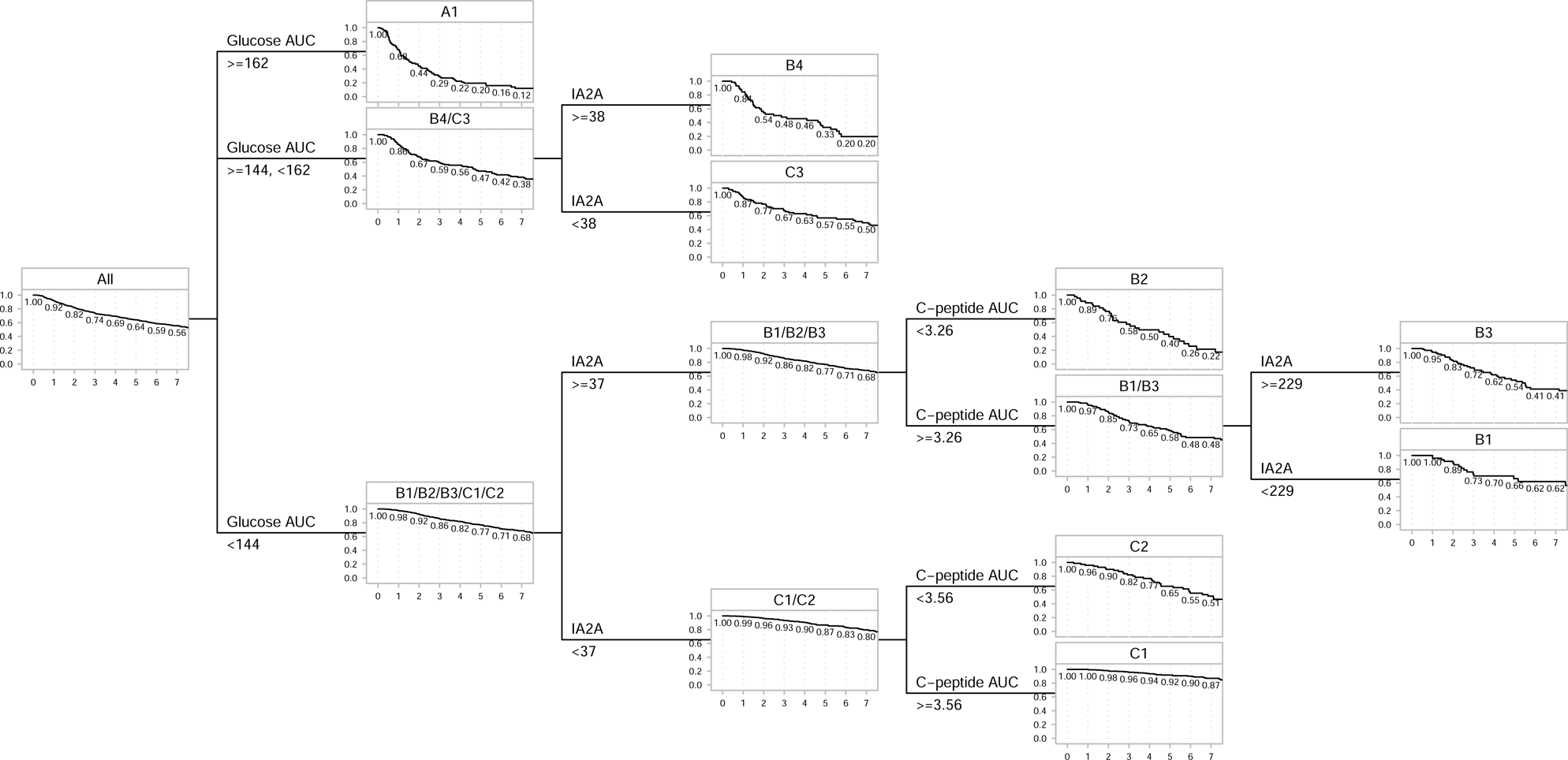
Decision rules to assign individuals to clusters.

As a validation of the results, we applied the decision rules to the 706 individuals in the DPT-1 dataset. The demographics of the individuals included in the validation data are given in the third column of Table 1. The distribution of variables by cluster is summarized in Supplementary Table S5. The comparison of the diabetes-free survival rate in the training and the validation dataset by cluster is shown in Figure 4. The two-year diabetes-free survival rate in the 8 clusters are A1: 0·54 [95% CI: 0·44-0·67], B1: 0·98 [95% CI: 0·94-1·00], B2: 0·86 [95% CI: 0·79-0·94], B3: 0·91 [95% CI: 0·85-0·98], B4: 0·65 [95% CI: 0·53-0·79], C1: 0·93 [95% CI: 0·89-0·97], C2: 0·95 [95% CI: 0·91-0·99], C3: 0·68 [95% CI: 0·56-0·82]. P-values from log-rank tests show that there were no significant differences in the survival rate in the two populations in the identified clusters except that the lowest risk cluster, Cluster A1, has a slightly lower diabetes-free survival rate in DPT-1 than in TrialNet PTP. This difference is likely attributed to the difference in study population, with DPT-1 having more participants with higher numbers of positive autoantibodies and higher autoantibody titers (see Table 1).

**Figure 4.**
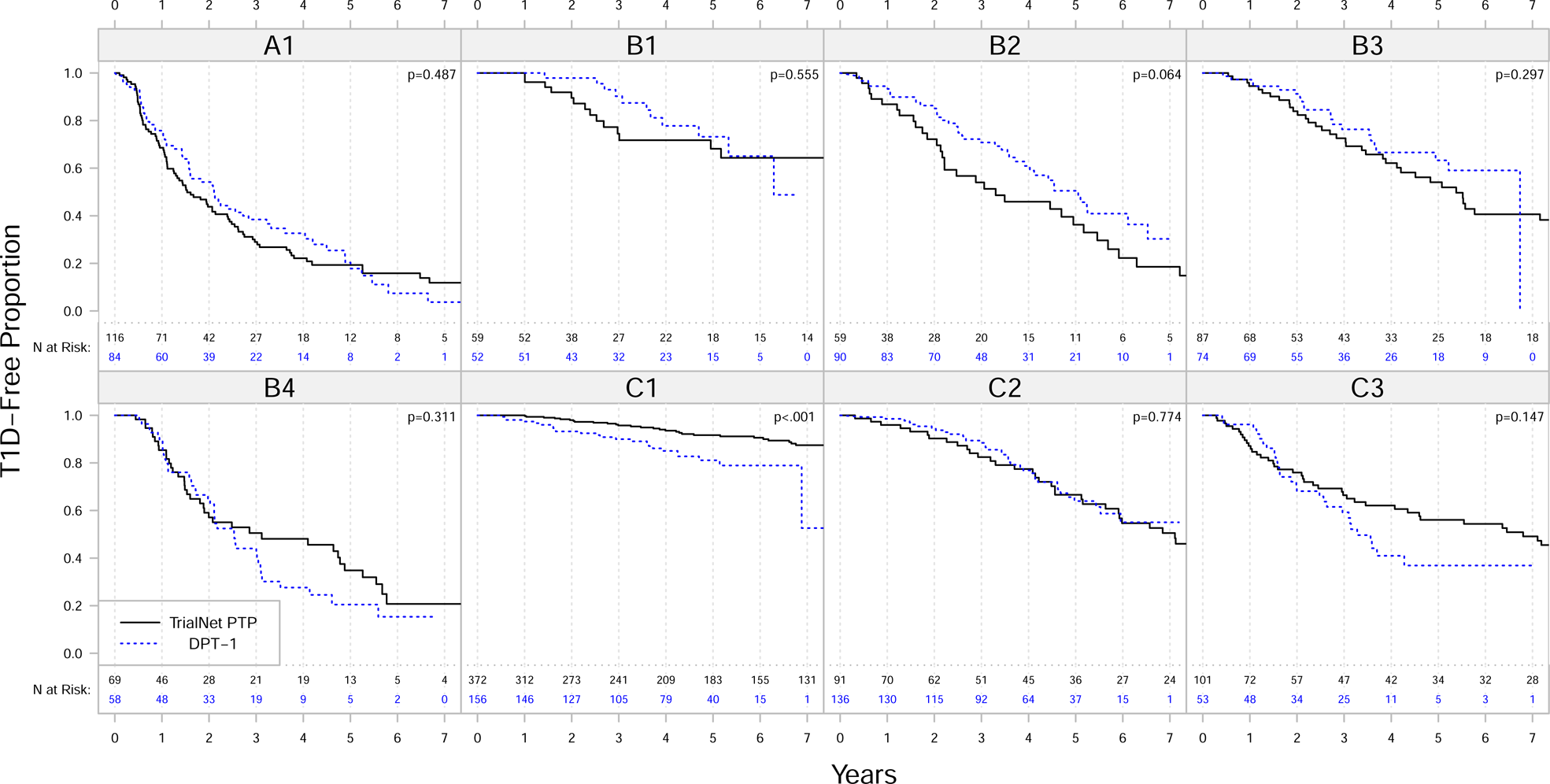
Comparison of diabetes-free Kaplan-Meier survival curves of clusters in the analysis dataset (TrialNet PTP; black solid lines) and the validation dataset (DPT-1; blue dashed lines). The numbers under the curves are the numbers of subjects at risk in the cluster at each time point P-values on the upper right corners are based on log-rank tests.

The comparison of variable distribution in the analysis dataset and validation dataset is presented in Supplementary Figure S5. Even though the two cohorts, TrialNet PTP and DPT-1 exhibit differences in variable distribution, there is an alignment in the distribution of variables by cluster, i.e., higher variable values in a cluster tend to align with higher values in the same cluster in the other cohort. For example, the DPT-1 population had higher BMI-z scores, higher GADA titers, lower mIAA titers, and higher ICA titers compared to the TrialNet PTP population. As a result, those variables are statistically different between the two cohorts for each of the clusters. However, similar to the relationships observed in TrialNet DPT, in DPT-1, cluster C1 still has a smaller proportion of individuals with positive autoantibodies, older ages and lower HbA1c levels. Cluster C2 identifies individuals with younger ages and a slightly higher rate of type 1 diabetes. Clusters B1, B2, B3 and B4 identifies individuals with larger numbers of positive autoantibodies and higher autoantibody titers. Cluster B1 and B3 have more individuals with older ages, B2 has more individuals with younger ages, and Cluster B4 has individuals with higher glucose levels. Clusters A1 consists of individuals with high glucose levels and higher HbA1c.

## Discussion

Metabolic, immunological and genetic markers can be used to identify clusters of distinctive characteristics and different risks of disease progression among islet autoantibody positive individuals with a family history of type 1 diabetes. Many risk factors that are known to predict disease were revealed by our analysis as variables for categorizing clusters. The clustering results demonstrate how different combinations and levels of these risk factors can contribute to the risk of type 1 diabetes, illustrating the heterogeneity of the population. Our approach ensures that the identified clusters are differentiated by their disease risk, while maintaining internal homogeneity within each cluster. As a result, the method can fulfill the goal of identifying clusters of individuals with similar risks but disparate disease risks across clusters. There is a parallelism between the identified clusters and the staging system for type 1 diabetes that is commonly used in the literature and clinical studies^25^. Cluster A consists of individuals with high glucose and HbA1c with elevated autoantibody titers and thus, there is a parallelism with stage 2 type 1 diabetes, i.e., dysglycemia with two or more positive islet autoantibodies, with a highly elevated risk of progression to stage 3 type 1 diabetes. Cluster B consists of participants with lower but still increased risk of type 1 diabetes, with elevated frequency of multiple autoantibodies (28·4%, 45·9%, 20·6%, and 5·1%, respectively, with four, three, two and single positive autoantibodies) and autoantibody titers (especially IA2A) but, compared with Cluster A, lower glucose levels. Therefore, Cluster B has similarities with Stage 1 type 1 diabetes, i.e., two or more islet autoantibodies with normal glucose tolerance. Finally, in line with previous findings that genetic susceptibility is the major risk stratification factor for pre-stage 1 type 1 diabetes, we observed that the lowest risk cluster (C1) has the lowest type 1 diabetes GRS2.

While there are similarities between the clustering scheme and the stages of type 1 diabetes when categorizing individuals by their glucose and autoantibody characteristics, this new method incorporates additional factors that interact with the key characteristics. For example, Cluster B2 is a high-risk cluster that cannot be identified by glycemic status alone as both glucose AUC and fasting glucose are low in this cluster. The elevated risk is primarily attributed to the presence of younger individuals with elevated autoantibody titers and genetic risk. Likewise, Cluster C2 includes younger individuals with lower glucose AUC, but the risk of type 1 diabetes in this cluster is elevated due to low C-peptide, high GRS2 and high IA2A titers.

From Figure 3, we can see that the decision tree for assigning individuals to clusters mainly relies on the three variables glucose AUC, C-peptide AUC, and IA2A titers. However, the clustering results show that the combination of the three variables is able to explain the differences we see in other variables. For example, in Cluster B2 which we characterize by glucose AUC <144, C-peptide AUC <3.26, and IA2A titer ≥137, the individuals have younger age, higher genetic risk, higher numbers of autoantibodies, and higher mIAA titers. In Cluster C1, which we characterize by glucose AUC <144, C-peptide AUC <3.56, and IA2A titer <137, there tend to be more individuals with older age and lower autoantibody titers of GADA, IAA and ICA as well. Furthermore, we can observe that clusters within the same branch but distinguished by a single variable can also identify distinct populations. For example, Clusters B4 and C3 are in the same branch with glucose AUC between 144 and 162 but differentiated by IA2A titers ≥38 vs <38. Individuals in Cluster B4 have higher numbers of autoantibodies and titers of IAA and ICA while individuals in Cluster C3 have older age, higher C-peptide levels and higher HbA1c. Our results suggest that GRS2 tends to be higher in Clusters B1, B2, and B3, and lower in Cluster C1 (Supplementary Table S4). However, GRS2 and genetic variants do not emerge as pivotal in defining the clusters. Additional research is required to investigate the connection between clusters and genetic factors.

In addition, this study shows that among individuals at similar risk of disease, there could be substantial differences in their metabolic and immunological characteristics. For example, we can compare Clusters B2 and C3 whose diabetes-free proportions are 0·87 vs 0·87 at 1 year, 0·72 vs 0·76 at 2 years, and 0·54 vs 0·66 at 3 years. The two clusters are similar in type 1 diabetes risk, but different in several aspects. Cluster B2 includes individuals with younger age, higher genetic risk, higher autoantibody titers, lower glucose levels, and lower C-peptide levels. In contrast, Cluster C3 includes individuals with older age, higher glucose levels, higher C-peptide levels, and higher HbA1c levels. Similarly, we can compare Clusters B1 and C2 whose diabetes-free proportions are 1.00 vs 0.96 at 1 year, 0.90 vs 0.90 at 2 years, and 0.74 vs 0.82 at 3 years. We can see that Cluster B1 has higher genetic risk, higher numbers of autoantibodies while Cluster C2 has younger age, lower HbA1c and lower numbers of autoantibodies.

There have been previous attempts to dissect the heterogeneity among individuals with pre-clinical type 1 diabetes. Our group previously observed that, in autoantibody-positive relatives who had normal 2-hour glucose in OGTT, Index60 (which combines C-peptide and glucose measures) could stratify participants with significant differences in age, autoantibody positivity, HLA associations and risk of progression to clinical (stage 3) type 1 diabetes^21^. In another study, it was found that among autoantibody positive relatives, individuals with a low Index60, regardless of their glucose levels, were older and more obese, and had a lower frequency of multiple autoantibody positivity compared to those with a higher Index60.^23^ Among patients with stage 3 type 1 diabetes, Taka et al.^26^ found similar heterogeneity in metabolic and immunological characteristics, with the presence of HLA-associated risk associated with multiple autoantibody positivity and younger age, while its absence was associated with diabetic ketoacidosis and older age. Overall, these results indicate that metabolic and immunological factors vary across the population and within groups of individuals with similar risk of progression, suggesting that there can possibly exist different disease pathways. The above observations also highlight another major difference between clustering analysis and prediction models. Prediction models assign everyone a risk score based on the individual’s characteristics while the clustering analysis will also reveal subgroups of individuals with similar characteristics within each risk stratum. Defining the trajectory of individuals as they progress will help characterize different pathways in the development of type 1 diabetes. This analysis will require using longitudinal data on the natural course of preclinical type 1 diabetes.

In this study, we also compared this method to previous prediction models based on multiple risk factors related to metabolic, immunological, and demographic characteristics. Most of the previous literature has used regression models to identify linear combinations of risk factors for disease prediction. For example, the DPTRS^6^ and the subsequent DPTRS60^7^ both use the proportional hazards model to define risk scores as linear combinations of age, BMI, and glucose and C-peptide measures from OGTT; DPTRS60 is a simplified version using only the first hour of OGTT results. Similarly, Index60 is a linear combination of OGTT-derived measure^7^. Bediaga et al.^9^ additionally included gender, IA2A titers, and HbA1c as risk factors in the model. Others have further considered expanding the list of predictors to incorporate genetic, demographic and other information.^8,27^ The risk scores derived from the regression models have been used to define risk groups by certain cutoffs. For example, a DPTRS threshold of 7·0 can reliably identify normoglycemic individuals at high risk^28^, and an Index60 threshold of 2·0 can result in an earlier diagnosis of type 1 diabetes compared to dysglycemia^7^. A combination of 2-hour glucose and Index60 can identify a subgroup of individuals at high risk of type 1 diabetes with younger ages, lower C-peptide, and multiple autoantibodies^23^. These prior efforts aimed to identify more homogenous subgroups of individuals for better prediction. However, prior methods using regression models assume that the risk increases linearly with the risk factors and neglect the interactions between risk factors that usually exist in a heterogeneous population. In contrast, our method is flexible to capture nonlinear structures when complex interactions exist.^13^ Although the regression methods can be improved to capture the interactions by adding interaction terms or subgroup analysis, the cutpoints for defining strata or subgroups are usually specified empirically by researchers or by evenly spaced quantile points^6,28,29^, while others also considered model-based approaches to search for an optimal cutpoint^27^. In contrast, the proposed tree-based clustering algorithm determines clusters and cutpoints by an automated algorithm. Additionally, Supplementary Figure S3 shows that overlaps between distributions of DPTRS and Index60 in different clusters exist and hence our results produce different separations of risk groups. For instance, Clusters C3 and C2 have similar DPTRS means, but different distributions of glucose AUC, C-peptide AUC, and age. Likewise, Clusters B2 and B4 have similar Index60 distributions, but Cluster B4 has a higher risk and higher glucose AUC compared Cluster B3 (Supplementary Figure S3).

There are several reasons why we chose the tree-based outcome-guided clustering method over other clustering and regression methods. As opposed to other methods, ours is outcome-guided, that is, it automatically selects variables that are predictive of progression to stage 3 type 1 diabetes to identify clusters. In addition, this method can easily handle datasets with a mixture of continuous, ordinal, and categorical variables. Compared to other clustering analyses in diabetes-related research, our proposed analysis is more robust and stable and optimizes the information gained from the dataset. Compared to the unsupervised clustering methods that identify clusters without reference to disease outcomes, the outcome-guided method we use defines clusters with respect to disease outcomes and tries to capture the clinically meaningful features in the data.

The clustering method that we present has limitations too. Our clustering method has limitations; for variables not in the decision tree, it is unclear how they affect disease risk simply based on the clustering results. The correlations between variables can also be obscured without further investigation. Also, we can observe that there still exists some unexplained heterogeneity within each cluster. For example, risk factors such as the type 1 diabetes GRS2 and the number of positive autoantibodies may additionally stratify Cluster C1; however, a larger sample size with more observed events can increase the number of clusters that can be robustly observed. In addition, the method assigns each individual to a discrete cluster, which can make it difficult to determine the impact of certain predictors on disease risk in the interpretation of results. The generalizability of the results is restricted by the population and variables that we selected. Finally, the analysis used participant characteristics only at baseline and thus cannot capture the longitudinal change and evolution of markers.

In sum, our analysis is unique in the literature as it describes differing features and their associated risks among autoantibody-positive individuals with familial predisposition for type 1 diabetes. The information can be integrated into prediction models and be used to identify the most beneficial subgroups for enrollment into prevention trials. Our analysis suggests that different pathways and mechanisms of disease progressions may exist. Assessment of the longitudinal pattern of risk factors would give more insights into disease staging and pathways.

## Data Availability

Clinical metadata and GRS genotyping data analyzed for this study are available in the NIDDK Central Repository at https://www.niddkrepository.org/studies/trialnet, in accordance with the NIDDK's controlled-access authorization process.

## Software Sharing Plan

The proposed method can be implemented using the R package “SurvivalClusteringTree” (https://cran.r-project.org/web/packages/SurvivalClusteringTree/index.html) developed by the authors.

## Acknowledgments

This research was funded by the National Institutes of Health (NIH) through the National Institute of Diabetes and Digestive and Kidney Diseases (R01DK121843). The Type 1 Diabetes TrialNet Study Group is a clinical trials network currently funded by the National Institutes of Health (NIH) through the National Institute of Diabetes and Digestive and Kidney Diseases, the National Institute of Allergy and Infectious Diseases, and The Eunice Kennedy Shriver National Institute of Child Health and Human Development, through the cooperative agreements U01 DK061010, U01 DK061034, U01 DK061042, U01 DK061058, U01 DK085461, U01 DK085465, U01 DK085466, U01 DK085476, U01 DK085499, U01 DK085509, U01 DK103180, U01 DK103153, U01 DK103266, U01 DK103282, U01 DK106984, U01 DK106994, U01 DK107013, U01 DK107014, UC4 DK106993, UC4DK117009, and the JDRF. The authors would like to acknowledge TrialNet participants, their families, research staff, and investigators who directly or indirectly contribute to the studies.

