## Supplementary Materials for "Type 1 Diabetes Risk Phenotypes Using Cluster Analysis"

**Supplementary Tables**

Supplementary Table S1. Type 1 Diabetes TrialNet Study Group

Personnel as of 4/28/2023.

Steering Committee: Kevan Herold (Yale University), Mark Anderson (University of California, San Francisco), Mark A. Atkinson (University of Florida), Todd Brusko (University of Florida), Jane Buckner (Benaroya Research Institute), Mark Clements (The Children’s Mercy Hospital), Peter G. Colman (Walter & Eliza Hall Institute of Medical Research), Mark Daniels (Children’s Hospital of Orange County), Linda DiMeglio (Indiana University), Carmella Evans-Molina (Indiana University), Jason Gaglia (Joslin Diabetes Center), Stephen E. Gitelman (University of California, San Francisco), Robin Goland (Columbia University), Peter Gottlieb (Barbara Davis Center for Childhood Diabetes), Michael Haller (University of Florida), Carla J. Greenbaum (Benaroya Research Institute), Martin Hessner (Medical College of Wisconsin), Jeffrey P. Krischer (University of South Florida), Megan Levings (University of British Columbia), Ingrid Libman (University of Pittsburgh), Peter Linsley (Benaroya Research Institute), Alice Long (Benaroya Research Institute), Sandra Lord (Benaroya Research Institute), Wayne Moore (The Children’s Mercy Hospital), Antoinette Moran (University of Minnesota), Andrew Muir (Emory Children’s Center), Priya Pralahad (Stanford University), William Russell (Vanderbilt Eskind Diabetes Clinic), Jennifer Sherr (Yale University), Lisa Spain (National Institute of Diabetes and Digestive and Kidney Diseases [NIDDK]), Andrea Steck (Barbara Davis Center for Childhood Diabetes), John Wentworth (Walter & Eliza Hall Institute of Medical Research), Diane Wherrett (University of Toronto), Perrin White (University of Texas Southwestern), Darrell M. Wilson (Stanford University), William Winter (University of Florida).

Past Members: Peter Antinozzi (Wake Forest University), David A. Baidal (University of Miami), Manuela Battaglia (San Raffaele University), Dorothy Becker (University of Pittsburgh), Penelope Bingley (University of Bristol), Emanuele Bosi (San Raffaele University), Richard Insel (JDRF), Thomas Kay (St. Vincent’s Institute of Medical Research), Mikael Knip (University of Helsinki), Åke Lernmark (Skane University Hospital), Yuk-Fun Liu (University of Bristol), Jennifer B. Marks (University of Miami), Jerry Palmer (University of Washington), Mark Peakman (King’s College), Louis Philipson (University of Chicago), Alberto Pugliese (University of Miami), Philip Raskin (University of Texas Southwestern), Maria Redondo (Baylor College of Medicine), Henry Rodriguez (University of South Florida Diabetes and Endocrinology Center), Bart Roep (Leiden University Medical Center), Desmond A. Schatz (University of Florida), Jay S. Skyler (University of Miami), Jay M. Sosenko (University of Miami), Jorma Toppari (Hospital District of Southwest Finland), Anette Ziegler (Technical University Munich).

Executive Committee: Kevan Herold, (Yale University), Linda DiMeglio (Indiana University), Carla J. Greenbaum (Benaroya Research Institute), Jeffrey P. Krischer (University of South Florida), Ellen Leschek (National Institute of Diabetes and Digestive and Kidney Diseases [NIDDK]), Lisa Spain (National Institute of Diabetes and Digestive and Kidney Diseases [NIDDK]).

Past Members: Katarzyna Bourcier (National Institute of Allergy and Infectious Diseases [NIAID]), Richard Insel (JDRF), John Ridge (National Institute of Allergy and Infectious Disease [NIAID]), Jay S. Skyler (University of Miami).

Chair’s Office: Kevan Herold, (Yale University), Lisa Rafkin (University of Miami).

Past Members: Carla J. Greenbaum (Benaroya Research Institute), Irene Santiago (University of Miami), Jay S. Skyler (University of Miami), Jay M. Sosenko (University of Miami).

TrialNet Coordinating Center (University of South Florida): Jeffrey P. Krischer, Brian Bundy, Michael Abbondondolo, Rajesh Adusumalli, Logan Alford, Matthew Boonstra, Jessica Conaty, David Cuthbertson, Julie Ford, Jennifer Garmeson, Veena Gowda, Cameron Hainline, Brian Hays, Kathleen Heyman, Christina Karges, Amy Kunz, Shu Liu, Kristin Maddox, Colleen Maguire, Margaret Moore, Sarah Muller, Melissa Murray, Johanna Nesbitt, Ryan O’Donnell, Melissa Parker, MJ Pereyra, Francisco Perez Laras, Aswani Raheja, Devon Rizzo, Ariana Rojas, Cintia Reichert, Lisa Steward, Michael Taylor, Roy Tamura, Dena Tewey, Elon Walker-Veras, Jianmei Wang, Melissa Wroble, Lili Wumser, Lu You, Kenneth Young.

Past Members: Timothy Adams, Darlene Amado, Ilma Asif, Jenna Bjellquist, Laura Bocchino, Cristina Burroughs, Mario Cleves, Meagan DeSalvatore, Christopher Eberhard, Steve Fiske, Susan Geyer, Courtney Henderson, Martha Henry, Belinda Hsiao, Amanda Kinderman, Beata-Gabriela Koziol, Lindsay Lane, Ashley Leinbach, Jennifer Lloyd, Jamie Malloy, Julie Martin, Cameron McNeill, Jessica Miller, Thuy Nguyen, Jodie Nunez, Nichole Reed, Amy Roberts, Kelly Sadler, Tina Stavros, Christine Sullivan, Megan V. Warnock, Keith Wood, Rebecca Wood, Ping Xu, Vanessa Yanek.

National Institute of Diabetes and Digestive and Kidney Diseases [NIDDK]: Ellen Leschek, Lisa Spain.

Data Safety and Monitoring Board: Emily Blumberg (University of Pennsylvania), Sean Aas (Georgetown University), Gerald Beck (Cleveland Clinic Foundation), Rose Gubitosi-Klug (Case Western Reserve University), Dennis Wallace (Retired).

Past Members: David Brillon (Cornell University), Lori Laffel (Joslin Diabetes Center), Robert Veatch (Georgetown University), Robert Vigersky (Medtronic).

Infectious Disease Safety Committee: Brett Loechelt (Children's National Medical Center), Lindsey Baden (Brigham and Women's Hospital), Peter Gottlieb (Barbara Davis Center for Childhood Diabetes), Michael Green (University of Pittsburgh), Ellen Leschek (National Institute of Diabetes and Digestive and Kidney Diseases [NIDDK]), Ingrid Libman (University of Pittsburgh), Adriana Weinberg (University of Colorado), John Wentworth (Walter & Eliza Hall Institute of Medical Research).

Past Members: Nora Bryant (Joslin Diabetes Center),Yuk-Fun Liu (University of Bristol).

Laboratory Directors: Michael Sheldon (Infinity BiologiX), Adriana Weinberg (University of Colorado), William Winter (University of Florida), Liping Yu (Barbara Davis Center for Childhood Diabetes).

Past Members: Santica Marcovina (University of Washington), Jerry P. Palmer (University of Washington), Jay Tischfield (Rutgers University).

TrialNet Clinical Network Hub (Benaroya Research Institute): Arielle Pagryzinski, Emily Batts, Danielle Tom, Catherine Nguyen, Chris Budy.

Past Members: Annie Schultz, Kristin Fitzpatrick, Randy Guerra, Melita Romasco, Annie Shultz, Mary Ramey, Michele Patience-Staal, Meghan Tobin, Diana Skye, Christopher Webb.

Active Personnel at Clinical Centers Participating in the TN01 Protocol:

Barbara Davis Center for Childhood Diabetes, Aurora, Colorado: Andrea K. Steck, Lexie Chesshir, Peter A. Gottlieb, Lisa Meyers, Aaron W. Michels, Marian Rewers, Kimber Simmons, Morgan Sooy, Fatima Tensun, Taylor Triolo, Leah Galvez Valencia, Paula Wadwa, Ruthie Williamson.

Benaroya Research Institute, Seattle, Washington: Carla J. Greenbaum, Jane H. Buckner, Sadiq El'Amin-White, Sandra Lord, Bao Ng, Mary Ramey, Michael Richter, Elaine Sachter, Corinna Tordillos, Dana VanBuecken, Kimberly Varner, Heather White, Nancy Wickstrom, Cassandra Williams, Alyssa Ylescupidez.

The Children's Hospital of Orange County: Amrit Bhangoo, Mark Daniels, Daina Dreimane, Mark Daniels, Marissa Erickson, Timothy Flannery, Nikta Forghani, Sarah Hu, Himala Kashmiri, Anabel Palencia, Christina Reh, Francoise Sutton, Heather Speer, Lien Trihn.

The Children's Mercy Hospital, Kansas City, Missouri: Wayne Moore, Fadi Al Muhaisen, Jennifer Boyd, Julia Broussard, Mark Clements, Aliza Elrod, Katelyn Evans, Max Feldt, Kelsee Halpin, Heather Harding, Jennifer James, Terri Luetjen, Ryan McDonough, Susan Mitchell, Tiffany Musick, Emily Paprocki, Rhiannon Pomerantz, Nikita Raje, Luis Sainz y Diaz, Britaney Spartz.

Columbia University, New York, New York: Robin Goland, Mone Anzai, Magdalena Bogun, Rachelle Gandica, Natasha Leibel, Jacqueline Lonier, James Pring, Nathan Schwab, Kristen Williams.

Emory Children's Center, Atlanta, Georgia: Andrew Muir, Amber Antich, Kristina Cossen, Eric Felner, Lynette Gonzalez, Wanda Sanchez, Catherine Simpson.

The Hospital for Sick Children, Toronto, Ontario: Diane K. Wherrett, Lesley Eisel, Sarah McGaugh, Rebecca Stochinsky, Mary Jo Ricci.

Indiana University, Indianapolis, Indiana: Linda A. DiMeglio, Carmella Evans-Molina, Jamie Felton, Heba M. Ismail, Megan Kirchner, Anna Neyman, Juan Sanchez, Corinne Parks-Schenck, Emily K. Sims, Maria Spall.

Stanford University, Stanford, California: Darrell M. Wilson, Bonnie Baker, Karen Barahona, Bruce Buckingham, Priya Prahalad, Trudy Esrey.

University of British Columbia, Kelowna, British Columbia: Constadina Panagiotopoulos, Daniel Metzger, Lauren Semkow.

University of California, San Francisco, California: Stephen Gitelman, Fatema Abdulhussein, Natalie Aceves, Mark Anderson, Julissa Cabrera, Hannah Chessner, Abby Cobb-Walch, Laura Dapkus, Aristides Diamant, Marysol Gonzales Granados, Tina Hu, Karen Ko, Janet Lee, Roger Long, Isabella Niu, Srinath Sanda, Caroline Schulmeister, Priya Srivastava, Lorraine Stiehl, Christine Torok, Rebecca Wesch, Jenise Wong, Kevin Yen.

University of Florida, Gainesville, Florida: Michael Haller, Mark Atkinson, Brittany Bruggeman, Todd M. Brusko, Miriam Cintron, Kristin Dayton, Timothy Foster, Jennifer Hosford, Laura Jacobsen, Sarah Peeling, Danielle Poulton, Desmond Schatz, Malinda Tran, William Winter.

University of Minnesota, Minneapolis, Minnesota: Antoinette Moran, Shannon Beasley, Melena Bellin, Jane Kennedy, Janice Leschyshyn, Brandon Nathan, Beth Pappenfus, Ihsan Rizky, Muna Sunni.

University of Pittsburgh, Pittsburgh, Pennsylvania: Ingrid Libman, Dorothy Becker, Kelli DeLallo, Christine March, Carly Shelleby, Frederico Toledo.

University of Texas Southwestern, Dallas, Texas: Perrin White, Abha Choudhary, Yasmin Dominguez, Philip Raskin, Serey Sao.

Vanderbilt Eskind Diabetes Clinic, Nashville, Tennessee: William Russell, Faith Brendle, Justin Gregory, Brenna Hammel, Jenny Leshko, Daniel Moore, Kimberly Rainer, Tyler Smith.

Walter and Eliza Hall Institute of Medical Research, Parkville, Victoria: Peter Colman, John M. Wentworth, Candice Breen, Marika Bjorasen, Spiros Fourlanos, Leonard Harrison, Felicity Healy, Leanne Redl.

Yale University, New Haven, Connecticut: Jennifer Sherr, Kevan Herold, Lori Carria, Jeanine May, William Tamborlane, Eileen Tichy, Stuart Weinzimer, Kate Weyman.

Supplementary Table S2. List of HLA alleles included in the analysis, separately from those used for T1D GRS2 computation.

| HLA Type | HLA Alleles | | | Count (Frequency) |
|---|---|---|---|---|
| | DRB1 | DQA1 | DQB1 | |
| S1 | 04:05 | 03:01 | 02:01 | 40 (3·30%) |
| S2 | 04:01 | 03:01 | 03:02 | 458 (37·82%) |
| S3 | 03:01 | 05:01 | 02:01 | 535 (44.18%) |
| S4 | 04:02 | 03:01 | 03:02 | 65 (5·37%) |
| S5 | 04:04 | 03:01 | 03:02 | 171 (14·12%) |

Supplementary Table S3. List of non-HLA SNPs included in the analysis, separately from those used for T1D GRS2 computation.

| Non-HLA SNP ID (Chromosome, Gene) | Count (Frequency) | | |
|---|---|---|---|
| | Minor Allele Homozygous | Heterozygous | Major Allele Homozygous |
| rs2476601 (1p13.2, PTPN22) | 33 (2·73%) | 310 (25·60%) | 868 (71·68%) |
| rs2816316 (1q31.2, RGS1) | 28 (2·31%) | 349 (28·82%) | 834 (68·87%) |
| rs10517086 (4p15.2, LOC105374540) | 118 (9·74%) | 501 (41·37%) | 592 (48·89%) |
| rs4948088 (7p12.1, COBL) | 1 (0·08%) | 77 (6·36%) | 1133 (93·56%) |
| rs1004446 (11p15.5, INS) | 128 (10·57%) | 537 (44·34%) | 546 (45·09%) |
| rs2292239 (12q13.2, ERBB3) | 166 (13·71%) | 573 (47·32%) | 472 (38·98%) |
| rs3184504 (12q24.12, SH2B3) | 340 (28·08%) | 552 (45·58%) | 319 (26·34%) |
| rs12708716 (16p13.13, CLEC16A) | 123 (10.16%) | 530 (43.77%) | 558 (46.08%) |

Supplementary Table S4. Distribution of variables summarized as median (IQR) in the eight identified clusters.

| Cluster | Age (years) | Type 1 Diabetes GRS2 | Glucose AUC (mg/dl) | C-peptide AUC (pmol/ml) | Fasting Glucose (mg/dl) | Fasting C-peptide (pmol/ml) |
|---|---|---|---|---|---|---|
| A1 | 11 (7-22) | 12.74 (11.28-13.75) | 172 (166-186)***↑ | 5.04 (3.59-7.13) | 93 (86-102)***↑ | 1.55 (0.88-2.12) |
| B1 | 12 (10-16) | 13.01 (11.91-14.07)*↑ | 123 (115-128)***↓ | 5.14 (4.38-6.83) | 88 (85-93) | 1.47 (1.08-1.92) |
| B2 | 6 (4-8)***↓ | 13.11 (11.65-14.25)**↑ | 123 (114-132)**↓ | 2.62 (2.18-2.91)***↓ | 82 (80-88)***↓ | 0.72 (0.56-0.93)***↓ |
| B3 | 12 (9-17) | 13.05 (11.97-13.92)**↑ | 124 (112-133)***↓ | 5.36 (4.28-6.49)*↑ | 88 (84-94) | 1.50 (1.19-2.09)*↑ |
| B4 | 10 (7-14) | 12.91 (11.50-13.86) | 150 (146-156)***↑ | 4.73 (3.77-6.96) | 90 (83-96) | 1.38 (0.90-1.94) |
| C1 | 13 (9-33)***↑ | 12.04 (10.44-13.56)***↓ | 119 (110-131)***↓ | 5.49 (4.52-6.76)***↑ | 90 (85-94) | 1.53 (1.22-1.92)***↑ |
| C2 | 6 (4-8)***↓ | 12.64 (10.68-13.64) | 120 (105-128)***↓ | 3.04 (2.52-3.30)***↓ | 86 (80-90)***↓ | 0.89 (0.66-1.02)***↓ |
| C3 | 14 (8-35)**↑ | 12.13 (10.70-13.90) | 153 (148-158)***↑ | 5.44 (3.90-7.43)**↑ | 94 (88-100)***↑ | 1.64 (1.14-2.10)**↑ |

* P<0.05, ** P<0.01, *** P<0.001, ↑ higher compared to the population, ↓ lower compared to the population.

Supplementary Table S4 (Continued). Distribution of variables summarized as median (IQR) in the eight identified clusters.

| Cluster | HbA1c | Num. AB | GADA (DK units/mL) | IA2A (DK units/mL) | IAA (index) | ICA (JDF units) |
|---|---|---|---|---|---|---|
| A1 | 5.3 (5.1-5.6)***↑ | 3.0 (2.0-3.0)***↑ | 339 (89-621) | 133 (1-269)***↑ | 0.0085 (0.0020-0.0262)*↑ | 40 (0-160)***↑ |
| B1 | 5.1 (4.9-5.3) | 3.0 (2.0-3.0)***↑ | 234 (47-538) | 140 (70-187)***↑ | 0.0040 (0.0020-0.0125) | 20 (0-160) |
| B2 | 5.1 (4.9-5.3) | 3.0 (2.0-4.0)***↑ | 157 (24-351)*↓ | 268 (135-314)***↑ | 0.0190 (0.0050-0.0580)***↑ | 80 (0-160)***↑ |
| B3 | 5.1 (4.9-5.3) | 3.0 (3.0-3.0)***↑ | 468 (143-644)**↑ | 299 (268-334)***↑ | 0.0050 (0.0015-0.0205) | 80 (40-320)***↑ |
| B4 | 5.2 (5.0-5.4) | 3.0 (2.0-4.0)***↑ | 333 (52-556) | 277 (223-324)***↑ | 0.0080 (0.0020-0.0350)*↑ | 120 (4-320)***↑ |
| C1 | 5.0 (4.9-5.2)***↓ | 1.0 (1.0-2.0)***↓ | 258 (5-559)*↓ | 0 (0-1)***↓ | 0.0030 (0.0010-0.0130)***↓ | 0 (0-10)***↓ |
| C2 | 5.0 (4.9-5.2)**↓ | 2.0 (1.0-2.0)*↓ | 280 (32-574) | 0 (0-1)***↓ | 0.0140 (0.0010-0.0430)**↑ | 0 (0-20)**↓ |
| C3 | 5.2 (5.0-5.4)*↑ | 2.0 (1.0-2.0)**↓ | 271 (24-543) | 0 (0-3)***↓ | 0.0040 (0.0010-0.0240) | 0 (0-20)*↓ |

* P<0.05, ** P<0.01, *** P<0.001, ↑ higher compared to the population, ↓ lower compared to the population.

Supplementary Table S5. Distribution of variables summarized as median (IQR) by clusters in the validation dataset (DPT-1). The distribution of variables in a cluster is compared to the population using Wilcoxon's rank sum tests.

| Cluster | Age | Glucose AUC (mg/dl) | C-peptide AUC (pmol/ml) | Fasting Glucose (mg/dl) | Fasting C-peptide (pmol/ml) |
|---|---|---|---|---|---|
| A1 | 12.59 (8.39-18.28) | 174 (165-183)***↑ | 3.61 (2.57-4.99) | 92 (85-98)***↑ | 0.90 (0.60-1.40) |
| B1 | 11.60 (8.57-13.93) | 126 (119-134) | 4.51 (4.04-5.19)***↑ | 87 (82-90) | 1.30 (0.97-1.60)***↑ |
| B2 | 8.08 (5.44-10.79)***↓ | 120 (106-132)***↓ | 2.51 (2.01-2.83)***↓ | 83 (77-89)***↓ | 0.60 (0.50-0.80)***↓ |
| B3 | 13.37 (11.11-15.82)***↑ | 117 (108-127)***↓ | 4.30 (3.65-5.37)***↑ | 86 (80-90) | 1.25 (0.92-1.50)***↑ |
| B4 | 10.27 (8.18-12.83) | 151 (148-156)***↑ | 3.51 (2.57-4.31) | 87 (81-92) | 0.80 (0.60-1.10) |
| C1 | 14.45 (10.14-21.90)***↑ | 121 (108-131)***↓ | 4.63 (4.01-5.59)***↑ | 86 (81-92) | 1.20 (0.90-1.70)***↑ |
| C2 | 8.42 (5.90-11.80)***↓ | 117 (104-129)***↓ | 2.69 (2.16-3.16)***↓ | 84 (78-88)***↓ | 0.60 (0.40-0.80)***↓ |
| C3 | 12.47 (8.52-16.62) | 152 (149-156)***↑ | 4.11 (3.09-5.22) | 88 (85-94)**↑ | 1.00 (0.50-1.40) |

* P<0.05, ** P<0.01, *** P<0.001, ↑ higher compared to the population, ↓ lower compared to the population.

Supplementary Table S3 (Continued). Distribution of variables summarized as median (IQR) by clusters in the validation dataset (DPT-1). The distribution of variables in a cluster is compared to the population using Wilcoxon's rank sum tests.

| Cluster | HbA1c (%) | Num. AB | GADA (DK units/ml) | IA2A (DK units/ml) | IAA (index) | ICA (JDF units) |
|---|---|---|---|---|---|---|
| A1 | 5.5 (5.2-5.8)*↑ | 3 (2-3)*↑ | 514 (246-676) | 115 (0-292)*↑ | -0.0020 (-0.0191-0.0180) | 160 (70-320)*↑ |
| B1 | 5.3 (5.0-5.5) | 3 (3-3)***↑ | 490 (121-635) | 138 (64-196)**↑ | -0.0010 (-0.0070-0.0084) | 80 (40-160) |
| B2 | 5.3 (5.0-5.6) | 3 (3-4)***↑ | 424 (126-655) | 279 (150-316)***↑ | 0.0070 (-0.0027-0.0200)***↑ | 160 (80-320)***↑ |
| B3 | 5.4 (5.2-5.7) | 3 (3-3)***↑ | 485 (173-723) | 317 (291-353)***↑ | 0.0010 (-0.0030-0.0145) | 160 (80-320)***↑ |
| B4 | 5.3 (5.0-5.7) | 3 (3-4)***↑ | 531 (267-728)*↑ | 301 (216-345)***↑ | 0.0015 (-0.0045-0.0170) | 320 (160-640)***↑ |
| C1 | 5.3 (5.1-5.6) | 2 (1-2)***↓ | 487 (1-764) | 0 (0-1)***↓ | 0.0007 (-0.0083-0.0070)*↓ | 40 (20-160)***↓ |
| C2 | 5.3 (5.0-5.5)**↓ | 2 (1-2)***↓ | 356 (1-654)**↓ | 0 (0-1)***↓ | 0.0010 (-0.0057-0.0100) | 40 (20-160)***↓ |
| C3 | 5.5 (5.2-5.7) | 2 (2-3) | 504 (90-663) | 0 (0-4)***↓ | -0.0010 (-0.0100-0.0134) | 80 (40-160) |

* P<0.05, ** P<0.01, *** P<0.001, ↑ higher compared to the population, ↓ lower compared to the population.

Supplementary Figure S1. CONSORT diagram of analyzed samples.

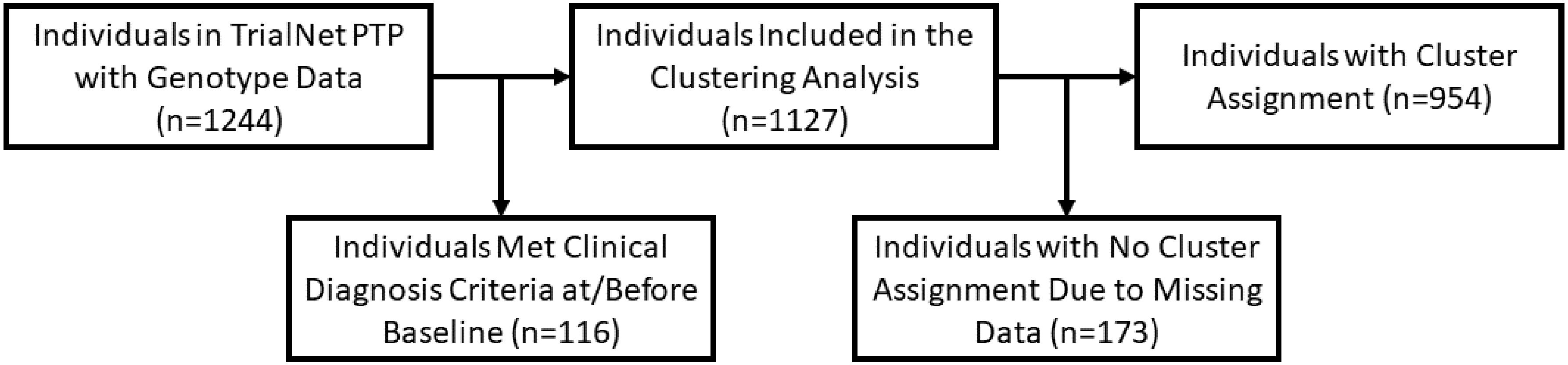

Supplementary Figure S2. A visual illustration of the outcome-guided clustering algorithm.

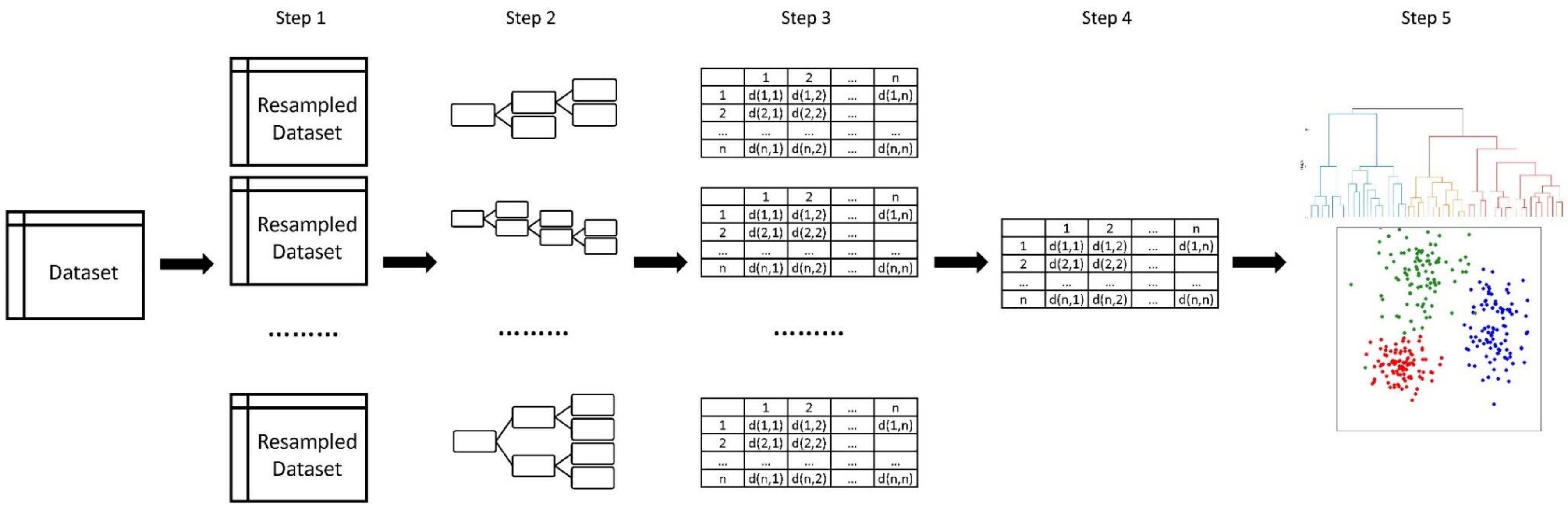

Supplementary Figure S3. Distribution of DPTRS and Index60 by cluster. Red bars in each cell represent the percentage of individuals in the DPTRS/Index60 intervals for each cluster.

| DPTRS | A1 | B1 | B2 | B3 | B4 | C1 | C2 | C3 |
|---|---|---|---|---|---|---|---|---|
| < 4.5 | 0.9% | 6.0% | 0.0% | 1.3% | 0.0% | 13.0% | 3.8% | 6.7% |
| [4.5,5.0) | 0.9% | 8.0% | 0.0% | 7.7% | 1.4% | 9.7% | 1.3% | 7.8% |
| [5.0,5.5) | 2.7% | 12.0% | 0.0% | 14.1% | 4.3% | 14.2% | 0.0% | 4.4% |
| [5.5,6.0) | 4.5% | 10.0% | 3.8% | 20.5% | 2.9% | 20.2% | 7.7% | 7.8% |
| [6.0,6.5) | 2.7% | 26.0% | 5.7% | 19.2% | 17.4% | 24.5% | 20.5% | 8.9% |
| [6.5,7.0) | 8.2% | 24.0% | 30.2% | 33.3% | 14.5% | 13.3% | 30.8% | 14.4% |
| [7.0,7.5) | 8.2% | 14.0% | 41.5% | 2.6% | 21.7% | 4.8% | 25.6% | 15.6% |
| [7.5,8.0) | 20.0% | 0.0% | 13.2% | 1.3% | 26.1% | 0.3% | 9.0% | 21.1% |
| [8.0,8.5) | 20.9% | 0.0% | 5.7% | 0.0% | 11.6% | 0.0% | 1.3% | 11.1% |
| 8.5+ | 30.9% | 0.0% | 0.0% | 0.0% | 0.0% | 0.0% | 0.0% | 2.2% |

| Index60 | A1 | B1 | B2 | B3 | B4 | C1 | C2 | C3 |
|---|---|---|---|---|---|---|---|---|
| < -2.0 | 1.7% | 5.1% | 0.0% | 1.1% | 0.0% | 3.8% | 0.0% | 5.0% |
| [-2.0,-1.5) | 2.6% | 1.7% | 0.0% | 4.6% | 1.4% | 3.0% | 0.0% | 4.0% |
| [-1.5,-1.0) | 3.5% | 10.2% | 0.0% | 6.9% | 7.2% | 7.5% | 0.0% | 5.9% |
| [-1.0,-0.5) | 2.6% | 13.6% | 0.0% | 10.3% | 4.3% | 21.2% | 0.0% | 6.9% |
| [-0.5,0.0) | 5.2% | 16.9% | 1.7% | 24.1% | 8.7% | 29.3% | 2.2% | 8.9% |
| [0.0,0.5) | 8.7% | 28.8% | 11.9% | 29.9% | 11.6% | 22.6% | 22.0% | 8.9% |
| [0.5,1.0) | 6.1% | 22.0% | 30.5% | 17.2% | 17.4% | 10.8% | 40.7% | 18.8% |
| [1.0,1.5) | 14.8% | 1.7% | 42.4% | 5.7% | 31.9% | 1.9% | 26.4% | 22.8% |
| [1.5,2.0) | 23.5% | 0.0% | 13.6% | 0.0% | 15.9% | 0.0% | 8.8% | 18.8% |
| 2.0+ | 31.3% | 0.0% | 0.0% | 0.0% | 1.4% | 0.0% | 0.0% | 0.0% |

Supplementary Figure S4. Measures for determining the number of clusters.

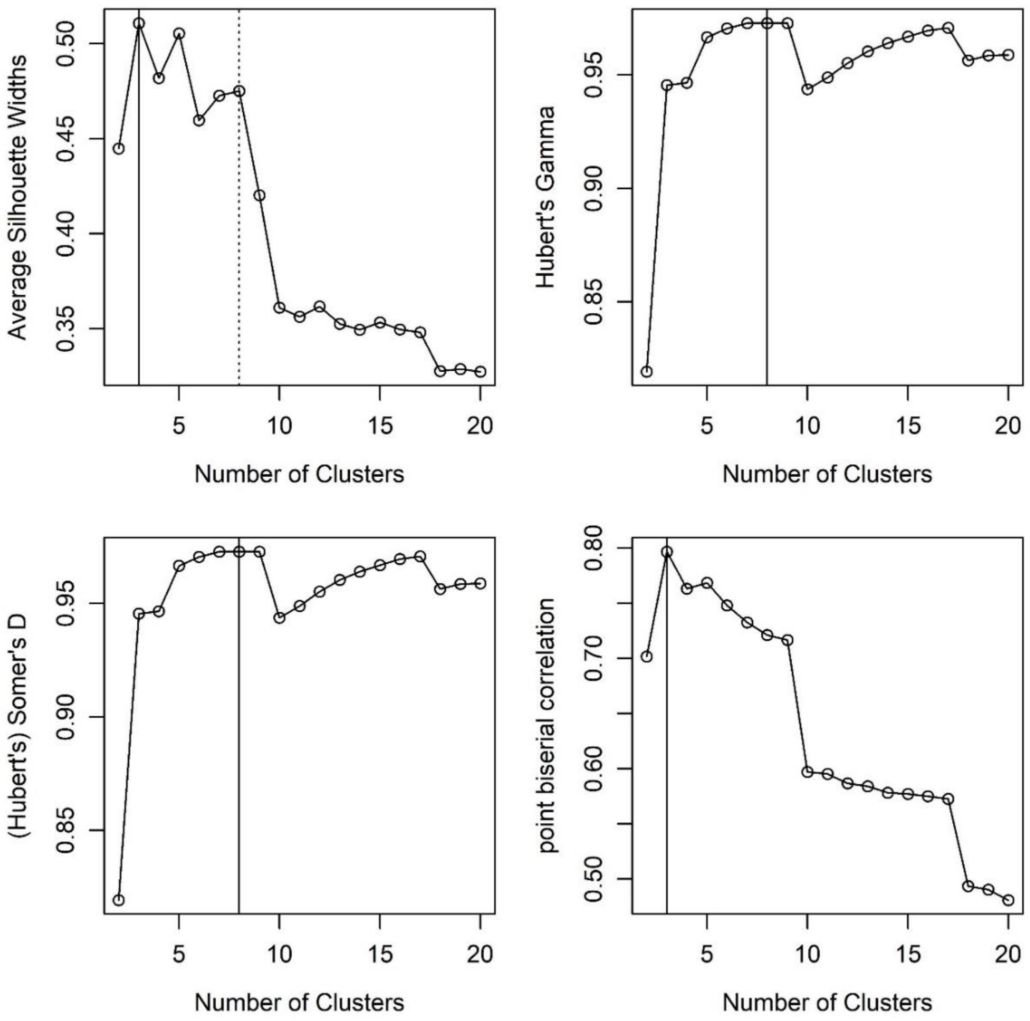

Supplementary Figure S5. Distribution of variables by clusters and the corresponding median (IQR) in the validation dataset (DPT-1) (blue bars) and the analysis dataset (TrialNet PTP) (orange bars).

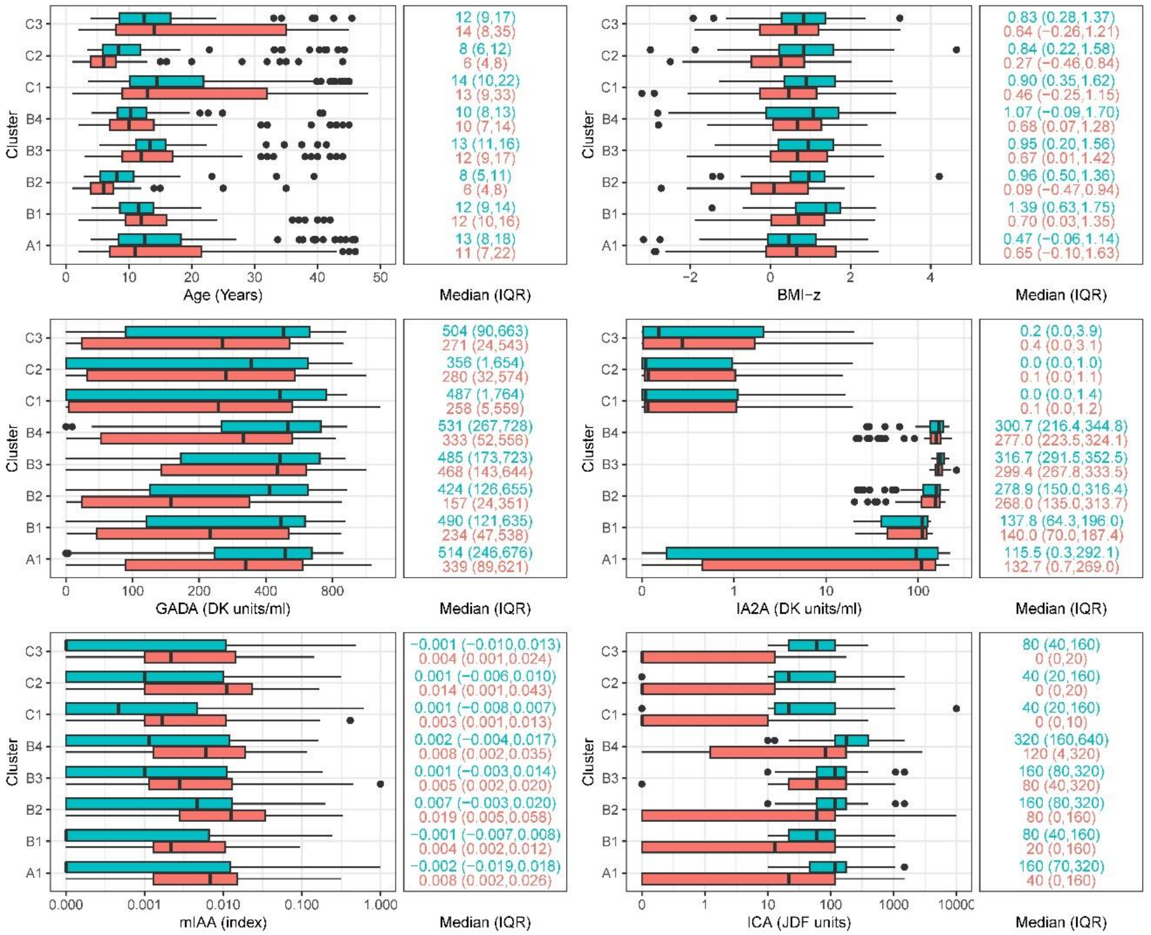

Supplementary Figure S5 (Continued). Distribution of variables by clusters and the corresponding median (IQR) in the validation dataset (DPT-1) (blue bars) and the analysis dataset (TrialNet PTP) (orange bars).

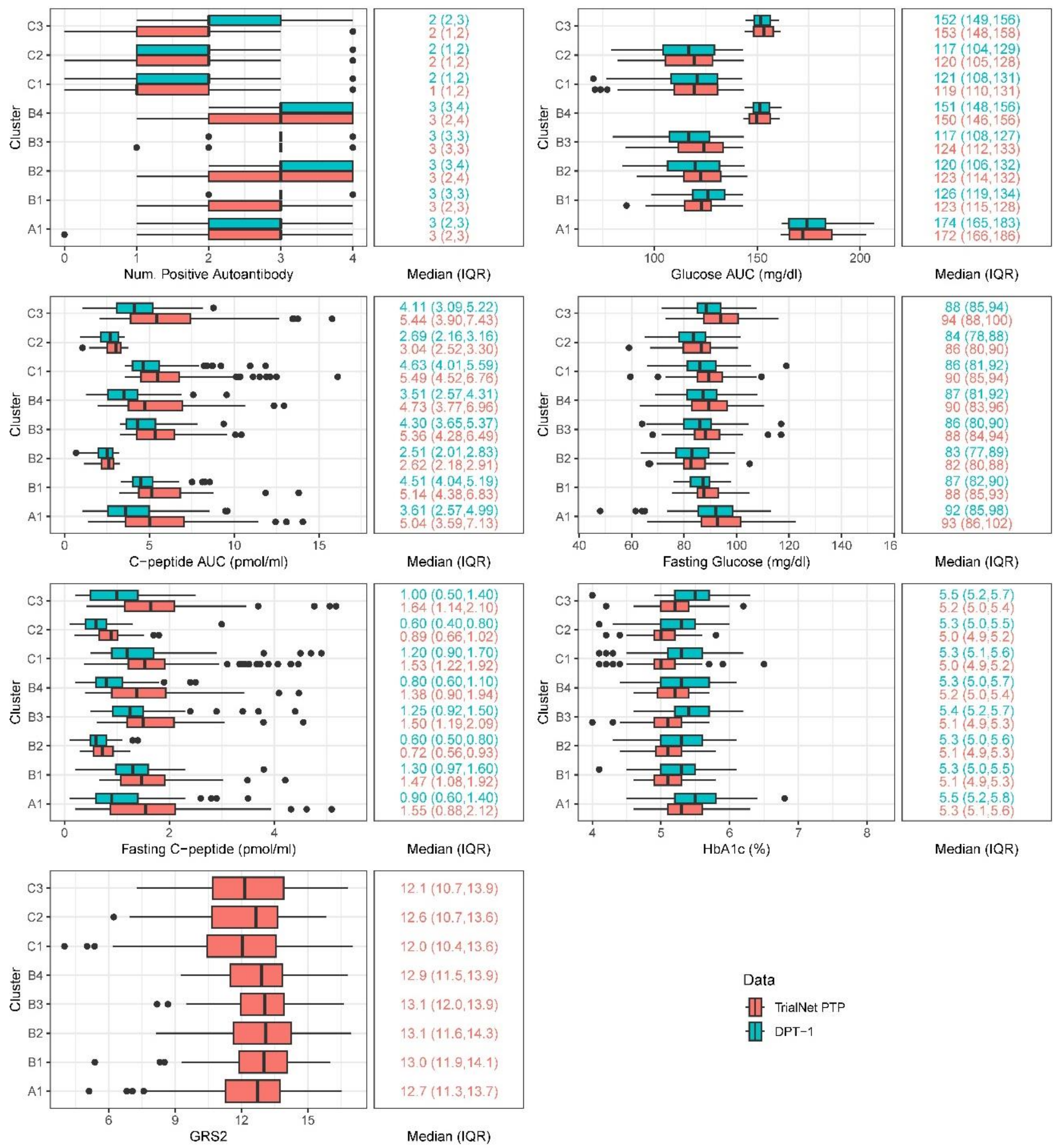
